# Key factors for effective implementation of healthcare workers support interventions in health organisations after patient safety incidents: a protocol for a scoping review

**DOI:** 10.1101/2022.01.25.22269846

**Authors:** Sofia Guerra Paiva, Maria João Lobão, João Diogo Simões, Helena Donato, José Joaquin Mira, Irene Carrillo, Paulo Sousa

## Abstract

**Introduction:** Health organisations should support healthcare workers who are physically and psychologically affected by patient safety incidents (second victims). There is a growing body of evidence which focusses on second victim support interventions. However, there is still limited research on the elements necessary to effectively implement and ensure the sustainability of these types of interventions. In this study, we propose to map and frame the key factors which underlie an effective implementation of healthcare worker support interventions in healthcare organisations when healthcare workers are physically and/or emotionally affected by patient safety incidents.

**Methods and analysis:** This scoping review will be guided by the established methodological Arksey and O’Malley framework, Levac and Joanna Briggs Institute (JBI) recommendations. We will follow the JBI three-step process: (1) a preliminary search conducted on two databases; (2) the definition of clear inclusion criteria and the creation of a list of search terms to be used in the subsequent running of the search on a larger number of databases; and (3) additional searches (cross-checking/cross-referencing of reference lists of eligible studies, hand-searching in target journals relevant to the topic, conference proceedings, institutional/organisational websites and networks repositories). We will undertake a comprehensive search strategy in relevant bibliographic databases (Pubmed/Medline, Embase, CINHAL, Web of Science, Scopus, PsycInfo, Epistemonikos, Scielo, Cochrane Library and Open Grey). We will use the Mixed Methods Appraisal Tool (MMAT) 2018 version for quality assessment of the eligible studies. Our scoping review will be guided by the Preferred Reporting Items for Systematic Reviews and Meta-Analysis extension for Scoping Reviews (PRISMA-ScR).

**Ethics and dissemination:** This study will not require ethical approval. Results of the scoping review will be published in a peer-review journal and findings will be presented in scientific conferences as well as in international forums and other relevant dissemination channels.

This protocol is registered on the Open Science Framework (www.osf.io): <u>10.17605/OSF.IO/WZSE9</u>

**Strengths and limitations of this study:**

- To our knowledge, this is the first scoping review to map and frame the different organisational, operational and contextual factors which underlie the implementation of health worker support programmes after the occurrence of a patient safety incident.
- This scoping review uses a rigorous and transparent method for mapping the available evidence.
- Given the broad focus of this scoping review, we don’t expect that eligible studies will show a direct relation between the key factors and the effectiveness of the implementation of the support interventions.
- For better interpretation of the results, we will provide a quality assessment of the included studies, although quality assessment is not mandatory to include in scoping reviews.
- We will involve key stakeholders as an additional source of information to complement the literature search.

## INTRODUCTION

Patient safety incidents affect patients’ lives at different levels and globally impact health organisations and their health care workers [1,2]. A patient safety incident is an unintended or unexpected healthcare event that either causes harm to one or more patients (adverse event) or that doesn’t reach a patient but poses a risk of harm (near miss) [1,3].

Patients suffering direct harm caused by a healthcare incident are the ‘first victims’ of an adverse event. Approximately 1 in 10 patients admitted to a hospital will suffer an adverse event, which represents 4% to 17% of hospital admissions [4,5], as will 4 in 10 patients in primary and outpatient healthcare [6]. According to a meta-analysis published in 2019, half of patient harm is preventable, which means that 6% to 12% of the prolonged, permanent disability or death identified in this meta-analysis could be prevented [7].

In 2000, Albert Wu [8] drew attention to the ‘second victim’ phenomenon in an editorial entitled ‘Medical error: the second victim. The doctor who makes the mistake needs help too’ which focused on the need to support health professionals who suffered from involvement in an adverse event. There is a growing body of evidence which focusses on understanding physical and psychological consequences experienced by healthcare professionals who are involved in a patient safety incident which causes serious or minor harm or even did not cause direct harm to the patient (near miss) [9,10]. Anxiety about future adverse events, depression, shame [9], frustration, repetitive/intrusive memories, extreme fatigue, sleep disturbances [11] and burnout [12] are some of the most frequent consequences reported in the literature. These emotional and physical consequences can impact professional performance and self-confidence [9], which can increase the likelihood of being involved in future adverse events [9,13].

Several studies have mentioned that health professionals feel unprotected and that they lack support after involvement in a healthcare incident [9,14]. Healthcare worker support practices are still underdeveloped and underused in healthcare institutions [9,14]. Although growing attention is being paid to the importance of cultivating supportive and non-judgmental environments, the blame culture continues to be one of the most frequent problems in health organisations [15]. This culture may negatively impact healthcare workers that are experiencing the second victim phenomenon [16].

Health organisations should be responsible for providing tools and training to support healthcare workers after an incident occurs during healthcare, thereby contributing to a learning environment and safer healthcare [17]. Most of the support strategies that have been internationally adopted are programmes and interventions [17]. The common goals of established support programmes are to reduce the psychological distress of second victims due to incidents and to foster coping strategies [2]. We can find different formats for support interventions/programmes, such as online programmes, peer support programmes, specific support tools and resources for healthcare providers [18,19,20].

Improvements in patient safety culture [22], resilience and adaptative capacity [23] are some outcomes related to these types of interventions in healthcare organisations.

According to the Donabedian model for measuring quality in medical care [24], outcomes depend on factors related to structure and processes.

Some recent studies have shown the influence of organisational structures, processes and outcomes on the effectiveness of research implementation turned into practice [25,26].

The described healthcare organisational characteristics were clearly aggregated by Yano [25] into organisational structures, organisational processes and organisational outcomes (summarised in Table 1).

**Table 1.** Organisational structures, processes and outcomes adapted from Yano [21]

|  |  |
| --- | --- |
| **Organisational Structures** | Some examples:<br>• Size of the organisational unit(s) – number of facilities, beds, providers<br>• Services – general and specialty services<br>• Staffing characteristics and integrated networks<br>• Leadership structure/authority<br>• Resource allocation<br>• Organisational culture<br>• Work environment/organisational climate |
| **Organisational Processes** | • Management processes – practice arrangements, managerial coordination of services and follow up<br>• Communication processes, procedures, quality of interactions<br>• Relationships – nature of roles and responsibilities, interpersonal styles |
| **Organisational Outcomes** | • Process quality measures<br>• Intermediate outcome measures<br>• Global health status measures<br>• Utilisation measures<br>• Workflow or efficiency measures<br>• Costs |

Organisational structures are focused on ‘static’ resources, which means that the resources are related to infrastructures, tools, equipment, healthcare services, units and the staffing level of the function for managing and delivering services [27].

Organisational processes, on the other hand, tend to be more mutable as they refer to organisational actions, procedures and service coordination. Buse, Mays and Walt referred to processes as the way in which policies are initiated, developed or formulated, negotiated, communicated, implemented and evaluated [28].

Organisational outcomes are described as performance measures and include practice-level measures of effectiveness. As mentioned by Yano [25], ‘organizational outcomes are distinct only insofar as they represent what the entire practice or institution would experience as a whole once implementation is complete and are thus inter-related to other evaluation activities’.

According to San Martín-Rodríguez, Beaulieu, D’Amour and Ferrada-Videla [26], organisational factors combine institutional characteristics as well as communication and coordination mechanisms. Factors in epidemiology are considered to be ‘events, characteristics, or other definable entities that have the potential to bring about a change in a health condition or other defined outcome’ [29].

In accordance with the health policy implementation process, an effective implementation includes a complex set of interrelationships: content, processes, context and actors [28].

Beyond all the aforementioned domains associated with the implementation of interventions, healthcare workers that participate in the interventions are considered to be important stakeholders for evidence-based implementation. Participant contributions can lead to more effective implementation and to the facilitation of practical applications [30] which, in turn, may contribute to the sustainability of the interventions

### Study rationale

There is a growing body of published evidence that is already focused on second victim support programmes and other types of support interventions in health organisations for healthcare workers who are physically and/or emotionally affected by patient safety incidents. A preliminary search related to this topic revealed two recently published scoping reviews [17,31], a systematic review [2] and a meta-analysis [32]. The main focus of the published studies has been to describe health professional support interventions and resources along with their benefits for second victims and challenges in their implementation processes. The studies also identify coping strategies used by second victims as well as peer support experiences.

Studies have shown the existence of barriers to the implementation of and concerns about the use of these types of support interventions or tools by healthcare teams (concerns about confidentiality of programmes, time investment, reluctance to show vulnerability and ask for help, limited awareness of the second victim phenomenon, lack of financial resources and a culture of blame) [2].

Although support interventions have demonstrated utility for health organisations, health worker wellbeing and patient safety, there is still limited research on the elements necessary for effective implementation of these types of interventions in order to overcome difficulties in the implementation process and ensure the sustainability of these initiatives in health organisations.

Our study is based on the knowledge from the implementation science and health policy implementation, and on the empirical recommendations and stated preferences from the participants involved in the evidence-based programs/interventions. We believe that this study will contribute to the future implementation of these types of interventions in health organisations.

### Objectives

In this scoping review, we propose to map and frame the key factors which underlie an effective implementation of healthcare workers support interventions in health organisations when healthcare workers are physically and/or emotionally affected by patient safety incidents.

Key factors will be organized in 5 domains: operational attributes, organisational and contextual factors, relevant actors and healthcare workers recommendations.

Our study was based on the knowledge from the implementation science and health policy implementation, and on the empirical recommendations and stated preferences from the participants involved in the evidence-based programs/interventions. We believe that this study will contribute to the future implementation of these types of interventions in health organisations.

## METHODS AND ANALYSIS

Our scoping review protocol is registered on the Open Science Framework (www.osf.io): DOI 10.17605/OSF.IO/2W46H

A scoping review is a valid and comprehensive approach with a rigorous and transparent method for mapping the evidence available in a specific area in order to clarify key characteristics or factors related to a concept [33,34].

The methodology of this scoping review will be guided by the established Arksey and O’Malley methodological framework [35] as enhanced by Levac et al. [36] and Joanna Briggs Institute (JBI) recommendations [34]. In this study, we will follow the six-stages of: (1) identifying the research question(s); (2) identifying relevant studies; (3) selecting studies; (4) charting the data; (5) collating, summarising and reporting the results; and (6) completing a consultation exercise.

This study will be guided by the Preferred Reporting Items for Systematic Reviews and Meta-Analysis extension for Scoping Reviews (PRISMA-ScR) to ensure transparency of the obtained results [37].

To develop this scoping review, a multidisciplinary team composed of healthcare workers, researchers, academics and a qualified librarian will be used.

We will now describe the different stages of this scoping review according to the Arksey and O’Malley methodological framework [35] for scoping reviews.

### Stage 1: Research questions

#### Main research question

What are the key factors for an effective implementation of healthcare worker support interventions in health organisations after the occurrence of patient safety incidents?

#### Secondary research questions

What are the organisational factors that contribute to an effective implementation of a second victim support programme/other types of support intervention in health organisations when healthcare workers are physically and/or emotionally affected by patient safety incidents?

What are the operational attributes of a second victim support programme/other types of support intervention in health organisations when healthcare workers are physically and/or emotionally affected by patient safety incidents?

Who are the most relevant actors in second victim support programmes/other types of support intervention in health organisations when healthcare workers are physically and/or emotionally affected by patient safety incidents?

What are the contextual factors for second victim support programmes/other types of support intervention in health organisations when healthcare workers are physically and/or emotionally affected by patient safety incidents?

### Stage 2: Identification of the relevant literature

#### Identification process

In this study, we will follow the JBI three-step process: (1) a preliminary search conducted on at least two databases; (2) the definition of clear inclusion criteria and the creation of a list of search terms to be used in the subsequent running of the search on a larger number of databases; (3) possible additional searches (cross-checking/cross-referencing of reference lists of potentially eligible studies, hand-searching in target journals relevant to the topic).

#### Preliminary literature search

The preliminary search strategy was based on a literature review previously developed to clarify our inclusion and exclusion criteria. Our search strategy structure was guided by the Population, Concept and Context (PCC) elements of the inclusion criteria recommended by JBI for scoping reviews [34]. We want to achieve as much sensitivity as possible in the scope of our research.

The preliminary literature search was tested in two databases that are widely used in the Health Sciences: Pubmed/ MEDLINE and Web of Science Core Collection.

For the search strategy, we used Mesh and natural language terms.

In table 2, we present the search strategy used in Pubmed/ MEDLINE and Web of Science Core Collection.

**Table 2.**
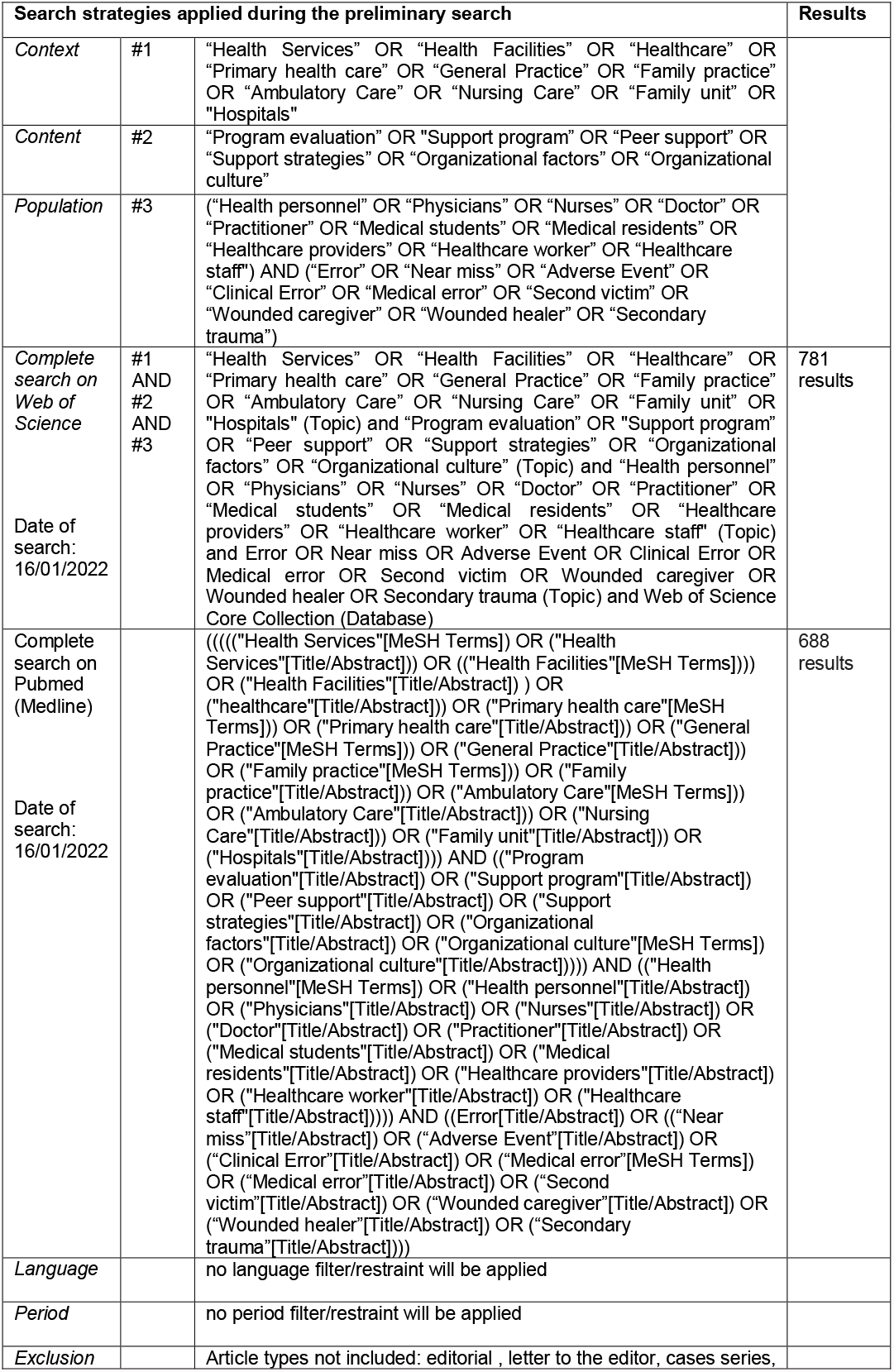
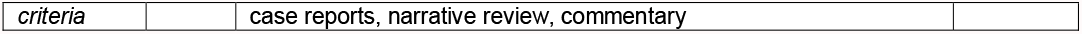
Preliminary literature search applied to Pubmed/ MEDLINE and Web of Science Core Collection databases

#### Structured search strategy

With the support of a qualified research librarian, we will undertake a comprehensive search strategy in relevant bibliographic databases, particularly in peer-reviewed journals and grey literature databases: Pubmed/Medline, Embase, CINHAL, Web of Science, Scopus, PsycInfo, Epistemonikos, Scielo, Cochrane Library and Open Grey.

We will also hand search in the reference list of the included articles, conference proceedings, institutional/organisational websites and networks repositories: The European Researchers’ Network Working on Second Victims (ERNST) website, Segundas y Terceras Víctimas Proyecto de Investigación, SARS-CoV-2 (COVID-19) Second Victims, Center for Patient Safety, Second Victim Support (UK) website, KU Leuven Research – Second Victim in Health Care, ForYOU team website, AHRQ website and World Health Organization (WHO) website.

We will conduct the searches from March to May 2022.

### Stage 3: Study selection

After applying the search strategy, results will be collated and exported to EndNote web.

Duplicates will automatically be removed for the further screening stages of study titles and abstracts of the eligible studies.

To reduce potential selection bias, studies will be independently screened by two authors. A third author can be involved in the case of disagreements.

To assure the maximum sensitivity of the screening process, the review team will do a pilot test with 30 randomly selected studies. We will screen this random sample using the agreed inclusion and exclusion criteria, discuss discrepancies and make any necessary modifications.

For full-text screening, we will note specific reasons for exclusion. Full texts will be included if at least two reviewers consider them eligible.

Screening results will be summarised and also graphically presented in a PRISMA flow chart (the number of excluded and included articles at each stage of the screening process will be reported).

#### Inclusion criteria

In our scoping review, we will consider the following criteria related to the typology of the studies:

- Study design/characteristics: original articles, systematic reviews and meta-analyses, grey literature (thesis, other documents);
- Period: no time filter/restraint will be applied; and
- Languages: no language filter/restraint will be applied.

We next describe the inclusion criteria according to Population, Concept and Context (PCC) elements.

##### Population

In this study, our target population is support interventions for healthcare workers who are involved in patient safety incidents.

The eligible interventions are focused on healthcare workers, defined as ‘people engaged in the promotion, protection, care or improvement of the health of population’, including health professionals and residents and other allied health professionals such as technicians and supply workers [38].

We will also include studies that use the term ‘second victims’, who are defined as ‘healthcare providers who are involved in an unanticipated adverse patient event, medical error and/or a patient related injury and become victimized in the sense that the provider is traumatized by the event’ [14]. However, there has been a discussion over the last few years about the adequacy of this term [39]. Therefore, in this scoping review, we will consider different terms for defining the second victim that have already been used in some of the literature, such as ‘secondary trauma’ [40], ‘wounded caregiver’ [41] and ‘wounded healer’ [42].

##### Concept

The concept of interest in this review is second victim support interventions/other types of support interventions in health organisations in which healthcare workers are physically and/or emotionally affected by patient safety incidents.

In this study, we intend to explore different types of healthcare worker support interventions: programmes, courses, pilot experiences and other tools and resources.

The development and/or the implementation and/or the evaluation of these types of support initiatives should be described in order to be eligible for this study.

We will focus on five key domains to study the implementation of these types of interventions (Figure 1).

**Figure 1:**
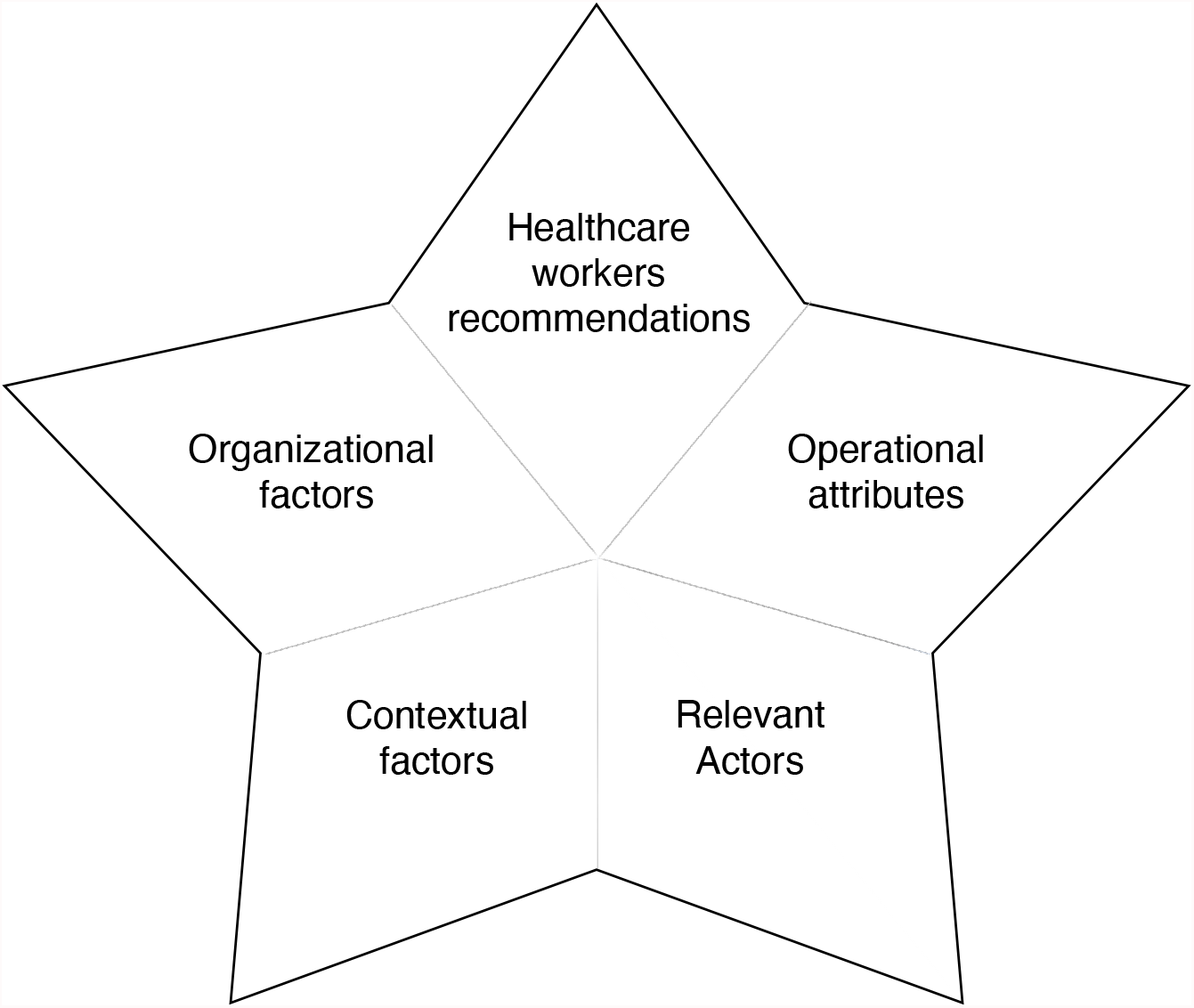
Five key domains for effective implementation of healthcare workers support interventions in health organisations

We will in particular consider the following aspects based on a health policy implementation and implementation sciences background:

- **Organisational factors**
  a. **organisational structures** such as infrastructures, resources, tools, equipment, units and staffing levels of function for managing and delivering services, leadership structure/authority and organisational culture;
  b. **organisational processes** such as organisational actions, procedures, recruitment criteria, training, programme implementation, communication processes and quality of interactions and coordination during programme implementation and dissemination as well as the sustainability of the practice;
  c. **organisational outcomes** such as implementation measures, process quality measures and utilisation measures; and effectiveness measures that assess the attainment of an end state, achievement of an objective, or creation of an effect in the healthcare organisation;
- **Operational attributes of the interventions** (format/type of programme, accessibility, usability and confidentiality of the programme/intervention);
- **Relevant actors** (the individuals and organisations that actively participate in the implementation);
- **Contextual factors** (type of healthcare setting and cultural context); and
- **Preferences and recommendations for the support programmes –** health care workers/second victim preference features and recommendations to improve the interventions.

##### Context

We will include all types of healthcare settings from high-, middle- and low-income countries in order to create a worldwide map of healthcare worker support interventions after the occurrence of a patient safety incident. According to the CDC Field Epidemiology Manual of the Center for Disease Control and Prevention, ‘healthcare settings represent a broad of services and places where healthcare occurs, including acute care hospitals, urgent care centers, rehabilitation centers, nursing homes and other long-term care facilities, specialized outpatient services (e.g., hemodialysis, dentistry,…), and outpatient surgery centers’ [43].

#### Exclusion criteria

In this study, we will exclude private offices and particular services due to their potential structural differences, specific contexts, and characteristics. Also, we will exclude the following types of publication: editorials, letters to the editor, cases series, case reports, narrative review, commentaries. A table of excluded evidence will be presented, with detailed reasons for their exclusion in the final scoping review

### Stage 4: Charting the data

This phase intends to give a logical and descriptive summary of eligible study characteristics and results. Arksey & O’Malley [35] referred to this phase as a ‘basic numerical analysis of the extent, nature and distribution of the studies included’. This will enable the alignment of study data with the objectives and questions of the scoping review [34].

A data extraction template will be created to show the characteristics of the eligible studies (table 3):

**Table 3.** Data extraction template for charting the data

|  |  |
| --- | --- |
| **Authors** | Study authors |
| **Year** | Year of publication |
| **Country** | Country where the programme was developed |
| **Population and setting** | Study population and setting where study was developed |
| **Aim/propose** | Aim/propose of the study |
| **Methodology** | Study design and types of methods used (survey, interview, observation, experiments...) |
| **Outcome measures** | Main results assessed in the study |
| **Key findings** | Results that relate to the scoping review questions |

The content of the template was developed by means of a preliminary exercise by the research team.

As recommended in the literature, a trial of the data extraction form was conducted by the reviewers with respect to at least three studies to ensure that all relevant information will be extracted from the eligible studies [34].

#### Quality assessment

There is an ongoing debate about the inclusion of an assessment phase related to the quality of eligible studies in scoping reviews [44]. Although this phase is not mandatory in this type of study [33,35], the absence of quality assessment is usually considered a methodological limitation. Some evidence shows that quality assessment can provide a better interpretation of results and an understanding of how the research was conducted, can clarify practice implications and can help to identify potential gaps in the evidence and the need for future research [33].

We will use the Mixed Methods Appraisal Tool (MMAT) 2018 version to conduct a quality assessment of the eligible studies. This is a validated tool to appraise the methodological quality of five categories of studies: qualitative research, randomised controlled trials, non-randomised controlled trials, quantitative descriptive studies and mixed methods studies [45]. Two independent reviewers will analyse the methodological quality of the included studies. A third reviewer will be involved in cases of disagreement in the quality assessment.

Information from each checklist item of the MMAT will be reported in a table format, described as ‘yes’, ‘no’ and ‘can’t tell’.

### Stage 5: Collating, summarising and reporting the results

After charting the data, each of the elements will be divided into different conceptual categories. Data will be collected in an Excel table using a descriptive content analysis. In addition, the results will be described in a narrative summary.

With respect to each of the included studies, we will extract the data as detailed in table 4.

**Table 4.** Data extraction template according to scoping review research questions

| Domains | Detailed description |
| --- | --- |
| Author, Year | Authors of the study, year of the study's publication |
| Type of intervention | Type of intervention described in the study (peer support programme, online programme, workshops, other) |
| Year of implementation | Year that support intervention was implemented |
| Duration of the intervention | Duration of the support intervention (in months) |
| Target population of the intervention | Health workers or medical residents/students (nurses, physicians, other allied health professionals such as technicians and supply workers,...) |
| Organisational factors | Some examples to consider in data extraction:<br>**a) organisational structures**<br>• Infrastructures (size of organisational or units, facilities, number of beds, ...)<br>• Resources, tools, equipment<br>• Staffing characteristics and integrated networks<br>• Leadership structure<br>• Organisational culture, work environment/organisational climate<br>**b) organisational processes**<br>• organisational procedures<br>• communication processes, interactions, roles and responsibilities<br>• management processes (practice arrangements, service coordination during programme implementation and follow-up)<br>• dissemination and sustainability of the practice<br>**c) organisational outcomes**<br>• process quality measures<br>• utilisation measures<br>• effectiveness measures<br>• global health status measures<br>• efficiency measures<br>• costs |
| **Operational attributes of the programmes/interventions** | Some examples to consider in data extraction:<br>• intervention type<br>• accessibility<br>• usability<br>• confidentiality issues<br>• other relevant attributes |
| **Relevant actors** | Individuals and organisations that make up and implement the programmes |
| **Contextual factors** | Type of healthcare setting, healthcare services and units, cultural context |
| **Healthcare workers/second victim recommendations** | Participant preference features and recommendations for the support intervention |

The content of the template was developed through a preliminary exercise by all the research team members. Data extraction can be updated during the data extraction process.

After collecting information in an Excel table (analytical framework), the information will be thematically organised according to the support intervention type provided to healthcare workers after the occurrence of an incident. In this phase, we will present information in a diagram using support intervention type as a primary unit of analysis.

### Stage 6: Consultation exercise and stakeholder involvement

A consultation exercise has been suggested in the literature [35,36] as an opportunity to involve key stakeholders as an additional source of information to complement the literature search. This will bring a high level of meaning content and will enlarge the scope of the review. In this phase, we will involve some of the members of the European Researchers Network Working on Second Victims (ERNST) - COST Action 19113.

## Data Availability

All data produced in the present work are contained in the manuscript

## Ethics and dissemination

This scoping review does not require ethical approval. Results and conclusions of the study will be published in a peer-reviewed journal and presented in scientific conferences as well as in international forums and through other relevant dissemination channels.

## Author contributions

SGP was involved in the writing of the article. SGP, MJL, PS and JJM contributed to the conceptualisation of the study. SGP, MJL and JDS analysed the data extraction templates, participated in the pilot testing stage and defined the inclusion criteria. HD contributed to the definition of a robust search strategy and participated in the preliminary search in relevant bibliographic databases, particularly in peer-reviewed journals and grey literature databases. PS, JJM and IC reviewed the article and contributed to the literature review. All authors read and approved the manuscript.

